# Surgical Resection, Radiotherapy, And Percutaneous Thermal Ablation for Treatment of Stage 1 Non-Small Cell Lung Cancer: A Systematic Review and Network Meta-Analysis

**DOI:** 10.1101/2021.09.20.21263867

**Authors:** Arun Chockalingam, Brandon Koo, John T. Moon, Menelaos Konstantinidis, Andrew Tran, Sahar Nourouzpour, Emily Lawson, Kathleen Fox, Peiman Habibollahi, Bruno C. Odisio, Mohammed Loya, Ali Bassir, Nariman Nezami

**Author notes:** **Corresponding Author:** Nariman Nezami, MD. 22 S. Greene Street, Baltimore, MD 21201.

## Abstract

**Introduction:** Non-small cell lung cancer (NSCLC) makes up the majority of lung cancer cases. Currently surgical resection of the affected lung parenchyma is the gold standard of treatment. However, as patients are becoming medically more complex and presenting with more advanced disease, minimally invasive image guided percutaneous ablations are gaining popularity. Therefore, comparison of surgical, ablative, and second-line external beam therapies will help clinicians, as management of NSCLC changes. We will conduct a meta-analysis, reviewing literature investigating these therapies in adult patients diagnosed with Stage I NSCLC (tumor ranging from 0-5 cm, with no hilar nor mediastinal nodal involvement, confirmed either through cytology or histology regardless of type).

**Methods and Analysis:** We will search electronic databases from their inception to January 2021 to identify randomized controlled trials (RCTs), cluster-RCTs, and cohort studies comparing the survival and clinical outcomes between any two interventions (lobectomy, wedge resection, radiofrequency ablation (RFA), microwave ablation (MWA), cryoablation and consolidated radiation therapies (EBRT, SBRT and 3D-CRT). The primary outcomes will include: cancer-specific survival (CSS), lung disease free survival, locoregional recurrence, death, toxicity, and non-target organ injury. In addition to the electronic databases, we will search for published and unpublished studies in trial registries and will review the references of included studies for possible inclusion in this review. Risk of bias will be assess using tools developed by the Cochrane collaboration. Two reviewers will independently assess the eligibility of studies and conduct the corresponding risk-of-bias assessments. For each outcome, given a sufficient number of studies, we will conduct a network meta-analysis. Finally, we will use the Confidence in Network meta-analysis (CINeMA) tool to assess the quality of the evidence for each of the primary outcomes.

**Ethics and Dissemination:** We aim to share our findings through high-impact peer review. As interventional techniques become more popular, it will be important for all providers in multi-disciplinary teams focused on care of these patients to receive continuing medical education on related to these interventions. Data synthesized in this study will be made available to readers.

## 1. BACKGROUND

### 1.1. Description of the Condition

Non-small cell lung cancer (NSCLC) accounts for 85% of all lung cancer cases and is a leading cause of cancer mortality worldwide. NSCLC is a heterogeneous set of epithelial lung malignancies consisting of three major subtypes: adenocarcinoma (ADC), squamous cell carcinoma (SCC), and large cell carcinoma (LCC). These subtypes together account for a 5-year survival rate of about 25% [1].

One factor that contributes to the poor survival is the nonspecific, late-appearing symptoms at presentation which cause delays in diagnosis. Approximately 75% of NSCLC cases present with inoperable locally advanced or metastatic disease, with a dismal 5-year survival rate of less than 5% [2].

Adoption of high-resolution low-dose CT (LDCT) screening with the intent of early detection in high-risk individuals has led to increased identification of smaller, early-stage NSCLC more amenable to curative intervention, with a correspondingly higher 5-year survival rate of 50-70% following resection. The advent of LDCT screening following the updated 2013 United States Preventive Services Task Force (USPTF) recommendations has also resulted in a stage-shift with increased incidence of stage 1 lung cancer cases from 19% to 27% [3]. The success in LDCT lung cancer screening has resulted in identification of a larger population of potentially curable stage I NSCLC. Surgical intervention (lobectomy, wedge resection) remains the gold standard for treatment of early-stage, resectable NSCLC. However, only 15-30% of patients are potential surgical candidates due to limited cardiopulmonary reserve, advanced age, and other disqualifying comorbidities [4]. This has led to an increased use of minimally invasive therapeutic options for high-risk patients with stage I NSCLC, including both radiation therapies and image-guided percutaneous ablation.

### 1.2. Description of the Intervention

Surgical resection has been the standard of care for early-stage NSCLC since the 1995 Lung Cancer Study Group Trial [5]. Surgical treatment in the form of lobectomy, or more limited sublobar resections (segmentectomy, wedge resection) involves the excision of lung parenchyma that is involved with tumor, leaving an appropriate margin of normal tissue. Anatomic resections, including lobectomy and segmentectomy, differ from wedge resection due to concurrent removal of interlobar and parenchymal (N1) lymph nodes for more accurate pathologic staging [6]. For patients with adequate physiologic reserve, lobectomy is preferred over wedge resection due to lower rates of loco-regional recurrence [5]. More recent studies since the 1995 LCSG trial have indicated equivalency in outcomes between lobar and sublobar resection, with no difference in long-term survival for stage I lung cancers [7, 8].

For the significant subset of older NSCLC patients with limited cardiopulmonary reserve who cannot tolerate surgery, a variety of non-surgical interventions including external beam therapies including stereotactic body radiation therapy (SBRT) or Proton therapy, and percutaneous thermal ablative therapies such as radiofrequency, microwave, or cryoablation have emerged as viable alternatives to surgical resection.

#### External beam therapies

##### 1. SBRT

SBRT, also referred to as stereotactic ablative radiotherapy (SABR), is a form of precision radiation therapy that delivers high doses of radiation to an image-defined mass. SBRT utilizes multiple beam angles to achieve sharp dose gradients that conform to the tumor with a minimal margin of surrounding normal tissue. SBRT also makes use of respiratory gating as well as patient immobilization systems to account for tumor motion during treatment delivery, allowing high dose radiation to be delivered to tumor in a reproducible manner [9]. Conventional radiation therapy for early-stage inoperable NSCLC has typically consisted of radiotherapy fractionated to a total dose of 60 Gy given over multiple fractions over several weeks [10]. In contrast, SBRT uses a hypofractionated regimen of five or fewer fractions of 10-20 Gy each to deliver the same total dose over a much shorter treatment period, reducing potential damage to surrounding tissue. In recent years, SBRT has emerged as the primary alternative therapy for early stage NSCLC to surgical resection, with an increase in utilization from 6.7% to 16.3% from 2008 to 2013, while the rate of lobectomy/pneumonectomy has declined from 49.5% to 43.7% over the same period [11].

##### 2. Proton therapy

Proton-beam therapy (PBT) is an evolving modality within radiotherapy involved with numerous cancer types, however its role in treatment of NSCLC is still not fully understood. As many as 25% of patients experience isolated locoregional recurrence and toxicity with standard radiation techniques [12]. PBT allows for sharp dose build-up and drop-off, reducing local exposure of radiation to uninvolved organs (i.e. heart, spinal cord, esophagus and healthy lung parenchyma). In addition, protons have a higher biologic effectiveness compared to photons [13], and as NSCLC patients live longer this becomes important to long term management of this disease. It is speculated that protons may spare circulating lymphocytes from radiation by reducing volume of blood that is radiated, preventing lymphopenia which has been correlated to worse outcomes [14]. However, proton therapy has been linked with increased chest wall pain and dermatitis, likely due to beam entry or end-ranging into thoracic chest wall anatomy [13] (Brooks, 2019). There is one randomized trial comparing protons to photons in early-stage NSCLC [15] which closed early due to concerns about lack of volumetric image-guided RT (IGRT) and poor accrual likely due to lack of insurance coverage. Comparison of this new form of radiotherapy to gold standard surgery and emerging interventional techniques would help assess efficacy and aid further investigation.

#### Percutaneous thermal ablative therapies

Using CT guidance, probes can be percutaneously inserted through the chest wall, and once an adequate ablative radius is imaged, the ablative therapy of choice (i.e. radiofrequency, microwave, and cryoablation) can be administered.

##### 1. Radiofrequency ablation (RFA)

RFA involves the delivery of a high-frequency electrical alternating current (approximately 375-480 kHz) leading to ion movement producing heat and protein denaturation. Temperatures up to 95 degrees C can be reached in about 15 minutes; however, efficacy can be limited by the heatsink phenomenon from nearby vessels and airways that can shunt heat away to subtherapeutic levels. Unlike microwave and cryoablation, RFA utilizes a single active probe at a time due to potential for electrical interference between probes.

##### 2. Microwave ablation (MWA)

MWA stimulates water molecules in tissue to create heat, which leads to immediate protein denaturation. Theoretically, MWA can generate higher temperatures in a larger volume in a shorter time than RFA, although results between these ablative therapies are comparable. Microwave ablation is favorable in ablation adjacent to large vessels and airways due to lower sensitivity to the heatsink phenomenon.

##### 3. Cryoablation

Cryoablation utilizes pressurized argon and helium to freeze tissue in contact with the ablation probes. Multiple freezing cycles are required as air in normal lung parenchyma limits the size of the ice-ball that forms. Upon thawing, intra-alveolar hemorrhage displaces the air, allowing for larger ice-balls to form, increasing necrosis zone size. Freezing and thawing cycles induce protein denaturation, membrane rupture, and ischemia within the zone of ablation. Complications of percutaneous ablative therapies include pneumothorax, hemorrhage, and local probe site infections, which were rare [16, 17].

### 1.3. Why it is important to do this review

The ability of LDCT lung cancer screening to detect an ever-increasing number of stage I NSCLC patients amenable to curative therapy warrants a re-examination of the current therapeutic options available to this population. Though the efficacy of surgical resection of local disease in these patients has long been demonstrated, a majority of patients (70%) are unable to tolerate surgery, thus requiring alternative treatment options. In this systematic review and network meta-analysis, we will evaluate the variety of non-surgical interventions that have evolved to treat stage I NSCLC, including, radiation therapies and percutaneous ablative therapies.

## 2. METHODS AND ANALYSIS

This protocol has followed the guidelines of, and is in compliance with the Preferred Reporting Items for Systematic Review and Meta-analysis Protocols (Supplementary Material) [18]. Any changes to this protocol will be delineated in the final article and will be reflected in an updated version of the PROSPERO registration. The methods section of this protocol is based on previously published protocols [19].

### 2.1. Criteria for considering studies for this review

#### 2.1.1. Types of Studies

In the review, we will include all Randomized controlled trials (RCTs), cluster-RCTs, and cohort studies comparing any two interventions (see below) against one another or against a placebo. We will not exclude studies on the basis of language, date of publication, or setting.

#### 2.1.2. Types of participants

We will include adult patients diagnosed with Stage I NSCLC (tumor ranging from 0-5 cm, with no hilar nor mediastinal nodal involvement). We will include patients whose status was confirmed either through cytology or histology without regard to the type of NSCLC (i.e., adenocarcinoma, squamous cell carcinoma, or large cell carcinoma).

#### 2.1.3. Types of interventions

We plan to compare 6 excisional, ablation, and radiation therapies. The excisional treatments are lobectomy, wedge resection. The ablation techniques are radiofrequency ablation (RFA), Microwave ablation (MWA), and cryoablation. Radiation therapies including EBRT, SBRT and 3D-CRT will be consolidated for ease of comparison. Given that this may induce some degree of heterogeneity, should we have sufficient studies, we will conduct our analysis with the more granular categorizations. Studies will be included if they compare any combination of interventions to one another or against no treatment or placebo. Should the specific treatment not be specified we will explore broader definitions of the interventions. As we are interested in the modality of the interventions, we will not define interventions in terms of the pharmacotherapeutic agents (where applicable); this will be explored as a subgroup analysis if possible.

#### 2.1.4. Types of outcome measures

##### 2.1.4.1. Primary Outcomes

⍰ Cancer-specific survival (CSS)
⍰ Lung disease free survival
⍰ Locoregional recurrence
⍰ Death
⍰ Toxicity
⍰ Non-target organ injury

##### 2.1.4.2. Secondary Outcomes

###### Major

⍰ Cancer-specific survival (CSS)
⍰ Disease-free survival (DFS)
⍰ Lung disease free survival
⍰ Residual disease
⍰ Locoregional recurrence
⍰ Toxicity
⍰ QALY
⍰ Infection
⍰ Hemoptysis
⍰ Bleeding
⍰ Hemothorax
⍰ Pneumothorax

###### Minor

⍰ Fever
⍰ Nausea
⍰ Vomiting
⍰ Shortness of breath
⍰ Chest pain

### 2.2. Search methods for identification of studies

We will conduct a systematic literature search to identify all published and unpublished trials for possible inclusion in this review. We will adapt the search strategy we developed for MEDLINE via PubMed (Supplementary Material) to search 5 electronic databases.

We will conduct searches in the following databases (from their inception to present):

- Embase (embase.com): 1947 – Present
- Web of Science (WoS) Classic Core Collections: 1900 – present
  - Includes: Science Citation Index Expanded, Social Sciences Citation Index, Arts & Humanities Citation Index, Conference Proceedings Citation Index-Science, Conference Proceedings Citation Index-Social Sciences & Humanities, Book Citation Index– Science, Book Citation Index– Social Sciences & Humanities, Emerging Sources Citation Index, Current Chemical Reactions, Index Chemicus
- Scopus (scopus.com): 1788 – Present
- Cochrane (cochrane.org): 1996 – Present
- ClinicalTrials.gov: 2008 – Present

We will apply no restriction on language of publications. We will also conduct a search of unpublished and ongoing trials in ClinicalTrials.gov (www.clinicaltrials.gov) (Appendix 6). In addition, we will screen the references of all included studies.

### 2.3. Data collection

#### 2.3.1. Selection of Studies

All references from the search strategy (see above) will be imported into COVIDENCE [20]. Two authors will independently screen abstracts and titles for possible inclusion in the review. The full text of the potentially included studies will be obtained, by which two authors will conduct a full-text assessment against the inclusion and exclusion criteria delineated above for inclusion of in the review, noting the reason for exclusion where applicable. At each stage, disagreement will be resolved through discussion, and where this is not possible, a third author will make a final decision. The final set of inclusions and exclusions will be depicted in a PRISMA flow chart [21].

#### 2.3.2. Data extraction and management

Two reviewers will independently extract the data from each of the included studies using a standardized extraction form (items enumerated below). As in the critical appraisal, any disagreements will be resolved through discussion or by a third reviewer if necessary. Relevant extracted data will be entered into Review Manager 5 [22] by one author and checked for accuracy by a second author. Data will then be exported to R [23].

From each study, insofar as the information is available, we will extract information on study characteristics, outcomes, and potential effect modifiers. If information is missing or additional information is needed (e.g., for risk of bias assessment), corresponding authors of the study will be contacted.

##### Study characteristics data

We will extract the article title, name of corresponding author, date of publication, type of publication, study setting, sources of funding, all conflicts of interest, study design, patient characteristics (e.g., age and gender), and study-level inclusion/exclusion criteria.

##### Outcome data

We will extract the interventions being compared and all primary and secondary outcome measures (and corresponding measure of uncertainty). In RCTs, we will aim to extract arm-level data or where this is not possible, study-level data. For observational studies, we will extract adjusted effect estimates, or if unavailable, unadjusted effect estimates – each with corresponding measure of uncertainty.

##### Potential Effect Modifier data

Additionally, we will extract trial size, rates of attrition. Moreover, if defined as an inclusion/exclusion criterion for a given study, we will extract the type of tumor, lung parity, node on which the tumor was found, and method of ascertainment for outcomes (only those corresponding to this review).

### 2.4. Risk of Bias

Results of the meta-analyses will be contextualized by the risk of bias from included studies. Two reviewers will independently assess the risk of bias of qualifying studies and as above, any disagreement will be resolved through discussion, or if necessary, with a third reviewer.

The Risk of Bias in RCTs will be evaluated using the Cochrane Risk of Bias (RoB) [24, 25] tool against the following domains:

⍰ Bias arising from the randomization process
⍰ Bias due to deviations from intended interventions
⍰ Bias due to missing outcome data
⍰ Bias in measurement of the outcome
⍰ Bias in the selection of the reported result

Based on signaling questions corresponding to each domain (“Yes”, “Potentially Yes”, “Potentially No”, “No”, or “No Information”), the risk of bias in each domain, as well as the overall risk of bias will be ranked as “low risk of bias”, “unclear risk of bias”, or “high risk of bias”. Should quasi-RCTs be included in the review, their corresponding risk of bias will be assessed using an additional domain as discussed in the *Cochrane Handbook for Systematic Reviews of Interventions* [25].

Non-randomized studies (NRS) will be evaluated using the Cochrane ROBINS-I tool – a tool that seeks to evaluate the risk of bias of a NRS as through the NRS were emulating an RCT (i.e., a target trial) [26]. The risk of bias will be evaluated across seven domains:

⍰ Bias due to confounding
⍰ Bias in selection of participants into the study
⍰ Bias in classification of interventions
⍰ Bias due to deviations from intended interventions
⍰ Bias due to missing data
⍰ Bias in measurement of the outcome
⍰ Bias in selection of the reported result

As in the case of the RoB tool, the bias for each domain, in addition to the overall risk of bias, will be assessed by series signalling questions (i.e., “Yes”, “Potentially Yes”, “Potentially No”, “No”, or “No Information”), yielding domain and overall risk-of-bias assessments of “low”, “moderate”, “serious”, or “critical”.

### 2.5. Data synthesis

#### 2.5.1. Characteristics of included studies

All included studies will be summarized through descriptive statistics. We will report on study population characteristics, including types of comparisons, design characteristics. For each outcome, we will present a network diagram, with nodes proportional to the number of patients, and edges proportional to the number of studies per comparison (Figure 1).

**Figure 1.**
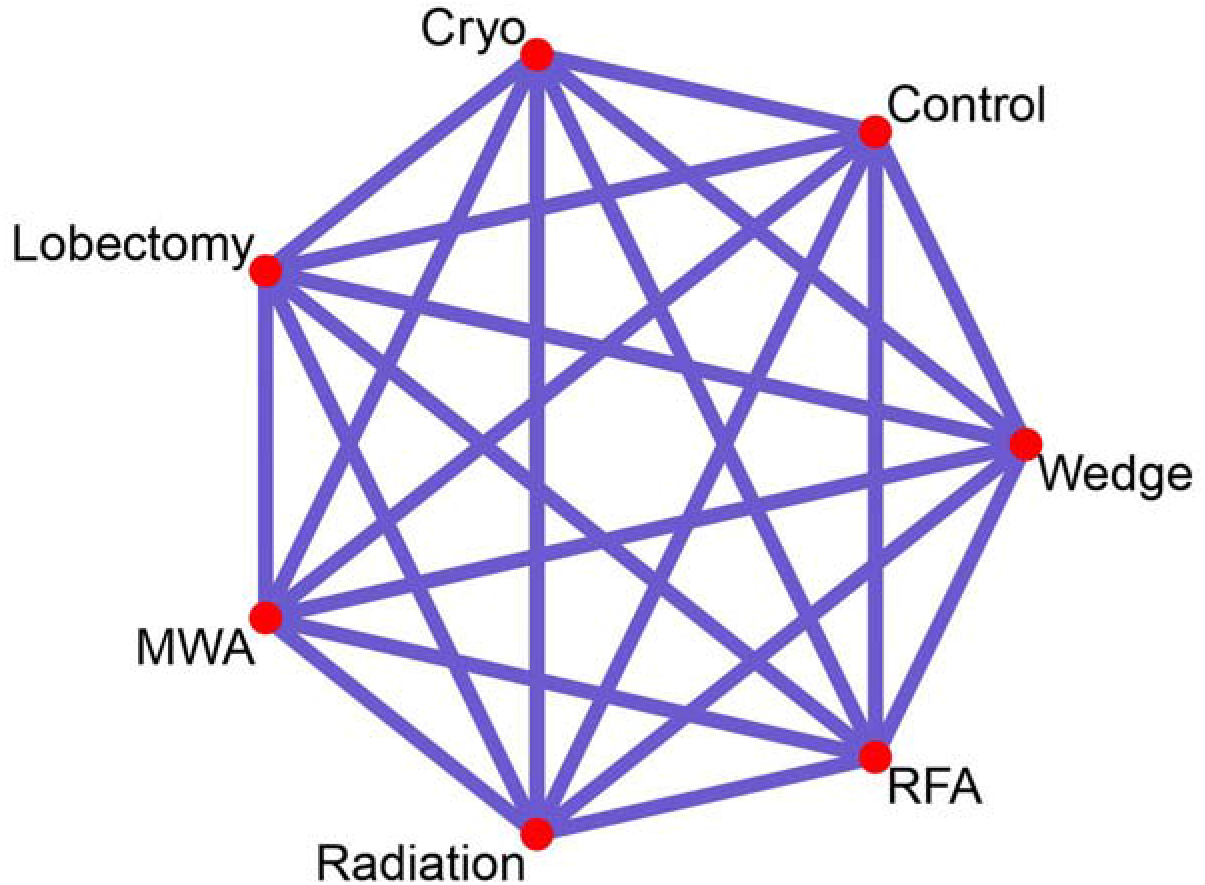
The network structure of all possible comparisons between Lobectomy, Wedge Resection (Wedge), Radiofrequency Ablation (RFA), Microwave Ablation (MWA), Cryoablation (Cryo), Radiation, and Control (e.g., no treatment). Red dots denote interventions and edges denote comparisons.

#### 2.5.2. Relative treatment effect

Pairwise relative treatment effects will be estimated using the odds Ratio for dichotomous outcomes, mean difference (MD) or standardized mean difference (SMD) for continuous outcomes, and hazard ratios for time-to-event or survival outcomes. For each outcome, we will report on the overall ranking of treatments obtained by the surface under the cumulative ranking curve (SUCRA) or P-Score providing a scalar value from 0 (least effective) to 1 (most effective) of each treatment for a given outcome [27].

#### 2.5.3. Pairwise meta-analysis

For each pairwise comparison, if more than one study is present, we will conduct a pairwise meta-analysis using an inverse-variance weighted random effects model. Estimation will be achieved using the restricted maximum likelihood estimator Between study heterogeneity will be estimated using the I^2^ statistic and corresponding 95% confidence interval [28]. Analyses will be carried out in R [23] [ref] using the meta package [29]. Given the possibility of small trials, we will estimate the summary effect estimates using the Hartung-Knapp-Sidik-Jonkman method [30, 31].

#### 2.5.4 Network meta-analysis

Given the likely clinical and methodological heterogeneity, we will fit a random effects network meta-analysis model for each outcome (data permitting) and will account for the correlation from multiarm trials. Common between-study variance will be assumed. Given that we will include both RCTs and non-randomized studies in the review, for each outcome, we will 1) fit a network to RCTs only, 2) fit a network to NRS only, 3) fit a network to all studies (RCT and NRS), and 4) fit a “design-adjusted” NMA, and 5) fit a “three-level hierarchical” NMA [32].

#### 2.5.5. Assessment of Heterogeneity

##### 2.5.5.1. Clinical and Methodological Heterogeneity

For pairwise meta-analyses, we will explore the heterogeneity by visual inspection of forest plots and estimated using the I^2^ statistic with corresponding 95% confidence interval [28, 33] [ref]. Additionally, we will pay attention to meaningful clinical groupings that may account for heterogeneity.

##### 2.5.5.2. Assessment of transitivity

A key assumption in NMA is that effect modifiers do not differ in their distributions across treatment comparisons. Thus, we will compare the distribution of the potential effect modifiers across each pairwise comparison using the direct evidence.

##### 2.5.5.3. Assessment of statistical inconsistency

To assess local inconsistency, we will use the back-calculation method [34]. In addition, we will use the design-by-treatment interaction model to assess global inconsistency [35]. Tests of consistency will be conducted in R using the netmeta package [36] [ref]. For networks with randomized and non-randomized studies, we will assess differences between the types of evidence (direct randomized, direct non-randomized, undirect randomized, and undirect non-randomized) as described in [32]. Data permitting, we will key discrepancies between the types of evidence will be further investigated, and if a source of disagreement is identified, where possible, we will conduct network meta-regression models [37].

#### 2.5.6. Unit of analysis issues and missing data

Multi-arm trials will be included and the correlation between effect estimates will be accounted for in the network. We do not anticipate any cluster- or crossover-RCTs. Should any such studies be included however, we will proceed under the guidance of the *Cochrane Handbook for Systematic Reviews of interventions* [25]. All analyses will be carried out on an intention-to-treat basis. For each included study, we will investigate the extent of missing data and evaluate the methods by which this was addressed in the risk of bias evaluation. For primary outcomes, we will conduct a sensitivity analysis to evaluate the impact of including studies for which there is a meaningful degree of missing data. Throughout, where possible, measures of uncertainty will be transformed to standard errors. Should no certainty be given or inferred, we will take the mean of known standard errors from the included studies [38, 39] [ref].

#### 2.5.7. Assessment of reporting biases

To assess the possibility of reporting bias and small-study effects, we will visually inspect the funnel plots [40] or each treatment where possible (i.e., at least ten studies for a given treatment). Additionally, to evaluate small-study effects, we will assess comparison-adjusted funnel plots [41], and where possible, conduct meta-regression with study variance as the covariate [42, 43].

### 2.6. Exploring heterogeneity and inconsistency

For the primary outcomes, we will explore the following sources of possible heterogeneity: Cancer type, lung parity, lobe on which node is found, and method of ascertainment for outcomes. Should sufficient studies be available, we will conduct subgroup analyses based on the characteristics. In addition, for the primary outcomes, we will endeavour to perform sensitivity analyses for:

- Study size (i.e., restricting to larger studies as smaller studies may lead to publication bias). Study size will be determined by the median of included studies, such that, in the sensitivity analysis, we will exclude studies whose size is below the median.
- Removal of studies with more than 20% missing data

### 2.7. Assessment of confidence in network estimates and summary of findings

The confidence of the network estimates for primary outcomes will be assessed using the Confidence in Network Meta-Analysis (CINeMA) framework [44, 45]. For each of the primary outcomes, two authors will independently assess the six CINeMA domains: within study bias, across study bias, indirectness, imprecision, heterogeneity, and incoherence [44] [ref] as very low, low, moderate, high confidence. This will be assessed using the online web tool [46].

## 3. Conclusions

### 3.1. Limitations

The proposed study is not without limitations. Given that we are using aggregated data for our planned meta-analyses as opposed to an individual-patient meta-analyses, we are only able to categorize our interventions into 6 categories: lobectomy, wedge resection, RFA, MWA, cryoablation, and radiation therapy. This means, however, that we will likely not be able to account for variations of a given treatment within or between studies. Additionally, since we are likely to compare multiple interventions, it is possible that we find statistically significant results by chance alone; however, there are no well-established methods to account for this issue in systematic reviews. With these considerations in mind, we believe that while valuable, the corresponding results should be interpreted with caution.

### 3.2. Summary

This systematic review and meta-analysis will serve as a foundation for comparison between the numerous treatment modalities and tool for clinicians to help best tailor therapies for early-stage NSCLC. As medicine advances, patients are presenting with more co-morbidities and may not be the best suited to receive surgery. Data assessing efficacy of interventional image-guided treatments compared to gold-standard will be crucial for evidence-based decision making in selecting therapeutic route and continuing innovation of these therapies.

### 3.3. Ethics and Dissemination

This study will not need new institution review board approval as this is a meta-analysis of previously published data. We aim to present our findings in high-impact peer-review journals and conference presentations. As interventional techniques become more popular, it becomes important for all clinicians involved in the care of these patients to receive continuing medical education on this topic. Data synthesized in this study will be available to readers.

### 3.4. Patient and public involvement

No patients, nor any members of the public were involved in the development of this protocol, nor will they be included in the completion or dissemination of the corresponding review and results.

## Data Availability

Data synthesized in this study will be made available to readers.

